# Differential Mechanisms of Storage Symptoms After Stroke: A Symptom Subtype and Lesion Network Analysis

**DOI:** 10.64898/2026.08.26.26361491

**Authors:** Zhaoxia Wang, Pei Dai, Zhike Yin, Sihao Liu, Qian Wang, Yuexiu Li, Changbin Liu, Chunchen Xiang, Zehui Li, Ran Liu, Yumei Zhang, Dawei Zang, Huixian Yu

## Abstract

**Background:** Storage symptoms after stroke—isolated urgency, urgency with frequency, and isolated frequency—are common but traditionally attributed to a single overactive bladder mechanism via suprapontine disinhibition. However, clinical heterogeneity in symptom presentation suggests distinct underlying mechanisms. We aimed to characterize the neural substrates of three storage symptom subtypes after stroke using comprehensive lesion-symptom mapping.

**Methods:** We prospectively evaluated 1,498 consecutive subacute stroke patients admitted for inpatient rehabilitation (1,105 men, 73.8%; median age 61 years). Storage symptoms were classified into three subtypes: isolated urgency (n=109), urgency with frequency (n=32), and isolated frequency (n=19). Multivariable logistic regression models with Bonferroni correction identified independent predictors across demographic, clinical, white matter hyperintensity (WMH), brain atrophy, and lesion location variables.

**Results:** The three subtypes demonstrated largely distinct sets of independent predictors. The left genu of the corpus callosum (aOR=20.06, 95% CI 7.78–51.74, *P*<0.001) and the inferior frontal gyrus (aOR=3.48, 95% CI 1.81–6.67, *P*<0.001) were independently associated with isolated urgency and survived Bonferroni correction, together with a right IFG–insula synergistic effect (OR=21.46, 95% CI 10.49–43.88, *P*<0.001). Urgency with frequency was associated with a broad fronto-cingulate network—the IFG (aOR=11.45, 95% CI 3.10–42.33, *P*<0.001, surviving Bonferroni correction) and the ACC (aOR=11.53, 95% CI 2.40–55.49, *P*=0.002)—with diffuse right-hemisphere dominance, older age and brain atrophy. Isolated frequency was associated with anterior corona radiata involvement (aOR=5.46, 95% CI 1.92–15.54, *P*=0.002) and male sex (aOR=10.62, 95% CI 1.36–82.98, *P*=0.024), though none reached the strict Bonferroni threshold.

**Conclusions:** These findings identify three mechanistically distinct post-stroke storage symptom subtypes with separable neural substrates, lateralization profiles, and clinical determinants. The triple dissociation across subtypes supports a discrete pathway model over the traditional unitary OAB framework, providing a neuroanatomically grounded basis for subtype-stratified treatment.

---

Post-stroke urinary incontinence is widely documented, yet prevalence estimates vary dramatically (15–60%), and identified risk factors remain inconsistent and often contradictory.^1,2^ Crucially, many studies conflate post-stroke incontinence with overactive bladder (OAB), a syndrome defined by urgency with or without UUI, usually accompanied by frequency and nocturia.^3,4^ Functional neuroimaging has provided critical insights into the central control of the bladder,^5^ and understanding of suprapontine pathways underlying voiding and storage is increasingly recognized as key to elucidating storage dysfunction.^6^ Although both conditions are classically attributed to suprapontine disinhibition, this unitary mechanism does not adequately explain why some patients present with urgency alone, others with urgency plus frequency, and still others with isolated frequency.^7.8^

In early-stage stroke survivors, storage symptoms manifest as distinct phenotypes that are likely to reflect divergent pathophysiologies, raising the possibility that each subtype originates from a distinct neural substrate. Forebrain control of urinary continence relies on a distributed cortical– subcortical network, and disruption at distinct levels of this hierarchy may yield discrete symptom profiles.^9^ Notably, Griffiths and Tadic demonstrated that cerebral responses to bladder filling consistently dissociate between patients with urgency urinary incontinence and those with detrusor overactivity, implicating aberrant bladder afferent signaling or impaired central processing thereof.¹⁰Building on these findings, Griffiths proposed a working model of the neural control of micturition,^10^ offering a mechanistic framework for how lesion location differentially compromises storage function. Convergent functional MRI evidence supports this view: the insula, a region central to interoception, is consistently engaged during bladder filling and urgency, suggesting that aberrant interoceptive processing contributes to storage dysfunction.^11^ Supraspinal fMRI studies in healthy men have further demonstrated that storage and micturition recruit dissociable supraspinal circuits,^12^ reinforcing the notion that damage to specific network nodes produces symptom-selective deficits. A particularly contentious issue is hemispheric laterality. Although functional imaging in non-stroke populations has repeatedly implicated the non-dominant (usually right) hemisphere—particularly the right inferior frontal cortex, a pivotal node of the interoceptive–salience network alongside the insula, in the inhibitory control and interoceptive perception of bladder filling,^13–15^ clinical urodynamic studies in stroke have yielded conflicting laterality findings.^8,13^ Whether laterality contributes to post-stroke storage phenotypes thus remains unresolved; prior work concluded that this correlation needs to be tested in a sufficiently powered lesion-symptom mapping study.^9^ Despite this accumulating evidence, no study has yet systematically distinguished these three storage symptom subtypes after stroke or examined whether they arise from distinct neural substrates. Resolving this heterogeneity would not only clarify the variable epidemiology of post-stroke incontinence but also offer a unique opportunity to refine our understanding of overactive bladder pathophysiology by linking specific symptom profiles to discrete lesion patterns.

Recruitment from a rehabilitation center offered a strategic advantage for investigating storage symptoms during the subacute phase of stroke. This temporal window is optimal for symptom ascertainment: the acute stage is often confounded by hemodynamic instability, fluid resuscitation, and polypharmacy, whereas in the subacute phase, stroke-related storage dysfunction has not yet undergone full compensation, rendering residual symptoms detectable and attributable to the lesion. Furthermore, early rehabilitation interventions in this setting mitigate confounders associated with prolonged bed rest and aphasia.

To address this gap, the present study had three aims: (1) to characterize the clinical and demographic factors associated with each storage symptom subtype after stroke (isolated urgency, urgency with frequency, and isolated frequency); (2) to map the neural substrates and hemispheric lateralization underlying each subtype using systematic lesion-symptom mapping, thereby directly addressing the unresolved question of laterality and generating neuroanatomically informed hypotheses regarding their separable mechanisms; and (3) to refine current models of overactive bladder pathophysiology by integrating these subtype-specific lesion signatures, ultimately providing a theoretical framework to guide personalized, lesion-targeted neuromodulation for storage symptom management in patients with both post-stroke and non-stroke etiologies.

## METHODS

### Study Population

The study protocol was approved by the institutional review board of the participating center (approval number: KY2021-035-02).

We prospectively enrolled 1,498 consecutive patients with subacute stroke admitted to the rehabilitation department between January 2022 and June 2026, all within 1 month of stroke onset. Inclusion criteria were: age ≥18 years; first-ever or recurrent stroke confirmed by CT or MRI; medical stability with urinary catheter removal before assessment; and the ability to cooperate with structured clinical assessment after cessation of significant intravenous fluid administration. Exclusion criteria were: persistent catheter dependence; functional incontinence secondary to severe cognitive impairment or mutism; and active urinary tract infection at the time of assessment (patients were treated and reassessed upon resolution).

### Clinical and Imaging Data

Demographic data (age, sex) were extracted from medical records. Stroke type was classified as ischemic or hemorrhagic. White matter hyperintensity (WMH) was graded using the Fazekas scale (0–6; positive ≥3).^16^ Brain atrophy was classified as present or absent from radiology reports. Functional status was assessed using the Barthel Index (BI); language function using the Western Aphasia Battery (WAB).

Lesion location, involvement of predefined structures, and laterality (left, right, bilateral) were determined from admission CT or MRI by a board-certified neuroradiologist and confirmed by consensus review. Analyzed structures included the anterior corona radiata (ACR), inferior frontal gyrus (IFG), insula, genu of the corpus callosum, anterior cingulate cortex (ACC), superior frontal gyrus (SFG), medial SFG, thalamus, basal ganglia, brainstem, and cerebellum.

### Lower Urinary Tract Symptom Classification

Lower urinary tract function was classified into four phenotypes from voiding diaries and structured assessment: (1) Normal: no new-onset urgency or frequency. (2) Isolated Urgency: sudden, unbearable urgency at normal bladder capacity, with or without urge incontinence, without significant frequency. In aphasic patients, urgency was inferred from non-verbal cues (restlessness, bed-tapping, gesturing to void). (3) Urgency with Frequency (U+Freq): urgency at small bladder capacity, accompanied by frequency (daytime ≥8 times and/or nocturia ≥2 times; voided volume <100 mL), excluding polyuria due to excessive fluid intake. (4) Isolated Frequency (FreqOnly): frequency meeting the above criteria, without urgency or urge incontinence.

### Statistical Analysis

Continuous variables were compared using Kruskal-Wallis tests with post hoc Mann-Whitney U pairwise comparisons. Categorical variables were compared using χ² or Fisher exact tests, with Cochran-Armitage trend tests for ordered groups. Univariate and multivariable logistic regression models estimated odds ratios (ORs) with 95% CIs for each subtype versus the normal group. Multivariable models simultaneously adjusted for age, sex, WMH, brain atrophy, and all lesion-location variables; the isolated-frequency model (n=19) included only IFG and ACR due to limited events. Bonferroni correction (α′=0.00152 for 3 subtypes × 11 variables) was applied; results reaching only nominal significance (P<0.05) are distinguished. Side-stratified analyses separately estimated left- and right-sided ORs. Synergistic effects were evaluated using exact 2×2 analyses with the relative excess risk due to interaction (RERI). Analyses were performed using Python 3.11 (SciPy, StatsModels). Two-tailed P<0.05 was considered significant.

## RESULTS

### Study Population

A total of 1,498 patients met inclusion criteria (1,105 men, 73.8%; median age 61 years, IQR 52–69), including 1,214 ischemic (81.0%) and 284 hemorrhagic (19.0%) strokes; median time from onset to assessment was 14 days (IQR 10–20), confirming the subacute phase. Of these, 160 (10.7%) reported storage symptoms: 109 isolated urgency (7.3%), 32 urgency with frequency (2.1%), and 19 isolated frequency (1.3%). Prevalence did not differ by stroke type (11.2% ischemic vs. 8.5% hemorrhagic, *P*=0.178).

### Clinical Profile

Patients with storage symptoms were older (median 68 vs. 61 years; *P*<0.001; OR per year 1.04, 95% CI 1.03–1.06) and had lower BI scores (median 40 vs. 55; OR per point 0.97, 95% CI 0.96– 0.98, *P*<0.001) than asymptomatic controls (Table 1, Figure 1A). They also showed higher odds of right-sided lesions (OR=1.76, *P*=0.001), bilateral lesions (OR=3.34, *P*<0.001), white matter hyperintensity (WMH; OR=2.03, *P*<0.001), and brain atrophy (OR=2.90, *P*<0.001). Aphasia was not significantly associated (OR=0.51, *P*=0.081), arguing against major ascertainment bias. Age increased across the ordered groups (Normal 61, Urgency 63, U+Freq 71, FreqOnly 72 years; Kruskal-Wallis *P*<0.001; trend β=4.11 years per group, *P*<0.001) and among the three storage-symptom subtypes alone (*P*=0.006).

**Figure 1.**
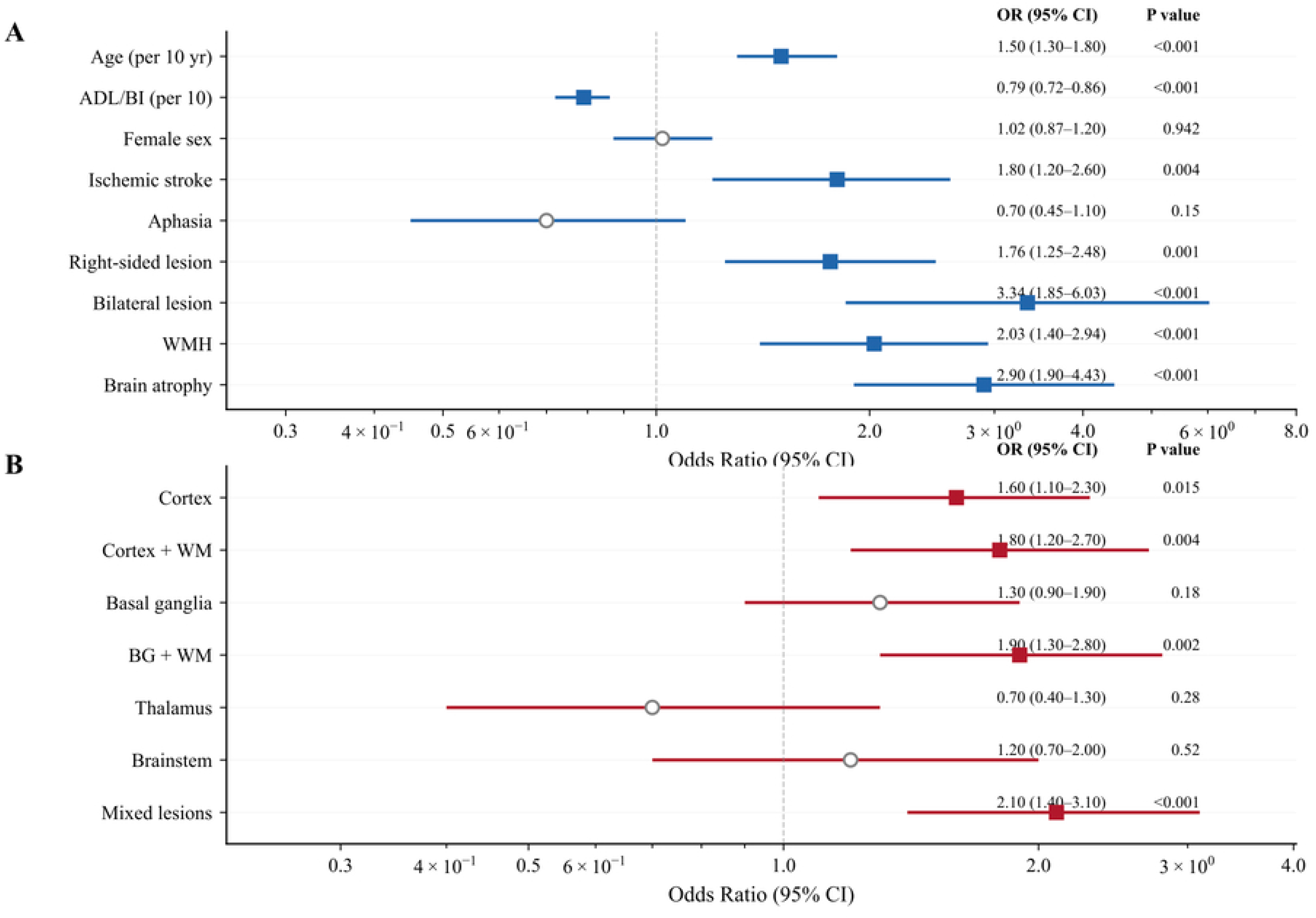
(A) Forest plot of clinical and demographic characteristics associated with storage symptoms (see Table 1). (B) Forest plot of lesion-location predictors from univariate logistic regression (see Table S1).

**Table 1.** Demographic and clinical characteristics of patients with and without post-stroke storage symptoms.

| Variable | Total<br>(n) | Normal<br>(n=1338) | Storage<br>Symptoms<br>(n=160) | Prevalence<br>(%) | OR<br>(95% CI) | P Value |
| --- | --- | --- | --- | --- | --- | --- |
| Age, median (IQR) |  | 61 (52–69) | 68 (60–73) |  | 1.04 (1.03–1.06) | <0.001 |
| ADL score, median<br>(IQR) |  | 55 (40–70) | 40 (30–55) |  | 0.97 (0.96–0.98) | <0.001 |
| Sex |  |  |  |  |  |  |
| Female | 393 | 357 | 36 | 9.2 | 1.00 (Ref.) |  |
| Male | 1105 | 981 | 124 | 11.2 | 1.25 (0.85–1.84) | 0.258 |
| Stroke type |  |  |  |  |  |  |
| Ischemic | 1214 | 1078 | 136 | 11.2 | 1.00 (Ref.) |  |
| Hemorrhagic | 284 | 260 | 24 | 8.5 | 1.36 (0.87–2.13) | 0.178 |
| Lesion laterality |  |  |  |  |  |  |
| Left | 778 | 717 | 61 | 7.8 | 1.00 (Ref.) |  |
| Right | 664 | 578 | 86 | 13.0 | 1.76 (1.24–2.49) | 0.001 |
| Bilateral | 59 | 46 | 13 | 22.0 | 3.34 (1.68–6.65) | <0.001 |
| Aphasia |  |  |  |  |  |  |
| No | 1365 | 1213 | 152 | 11.1 | 1.00 (Ref.) |  |
| Yes | 133 | 125 | 8 | 6.0 | 0.51 (0.25–1.07) | 0.081 |
| White matter<br>hyperintensity |  |  |  |  |  |  |
| No (Fazekas 0–2) | 752 | 689 | 63 | 8.4 | 1.00 (Ref.) |  |
| Yes (Fazekas ≥3) | 473 | 399 | 74 | 15.6 | 2.03 (1.42–2.89) | **<0.001** |
| Brain atrophy |  |  |  |  |  |  |
| No | 1310 | 1192 | 118 | 9.0 | 1.00 (Ref.) |  |
| Yes | 188 | 146 | 42 | 22.3 | 2.90 (2.01–4.20) | **<0.001** |

\*Abbreviations: ADL, activities of daily living (Barthel Index); CI, confidence interval; IQR, interquartile range.
†Data are median (IQR) or n (%); P from $\chi^2$ , Fisher exact, or Mann-Whitney U test. Bold indicates P<0.05.
‡Storage symptoms include urgency (with or without urge incontinence) and/or frequency. §Laterality counts sum >1498 (3 bilateral coded in >1 category). WMHs ungraded in 273 (18%).

Cochran-Armitage trend tests across ordered groups (Normal→Urgency→U+Freq→FreqOnly) revealed distinct patterns. The anterior corona radiata (ACR) showed a strictly monotonic increase (20.7%→36.7%→40.6%→57.9%; Z=5.76, *P*<0.001). Most other lesions peaked in the U+Freq group: WMH (peak 59.4%; Z=4.50, *P*<0.001), brain atrophy (peak 31.2%; Z=5.18, *P*<0.001), IFG (peak 37.5%; Z=7.17, *P*<0.001), ACC (peak 18.8%; Z=7.74, *P*<0.001), CC genu (peak 18.3% in Urgency; Z=6.93, *P*<0.001), and cortical involvement (peak 65.1% in Urgency; Z=5.29, *P*<0.001). Aphasia decreased across subtypes (9.3%→7.3%→0%→0%; Z=−2.26, *P*=0.024).

### Symptom Subtype-Specific Multivariable Predictors

Multivariable logistic regression revealed largely distinct predictor sets for the three subtypes (Figure 2), though few reached the Bonferroni threshold (α′=0.00152). For isolated urgency, the CC genu (aOR=20.06, 95% CI 7.78–51.74, *P*<0.001) and IFG (aOR=3.48, 95% CI 1.81–6.67, *P*<0.001) survived correction; WMH (aOR=2.35, *P*=0.004) and BI (aOR=0.97, *P*<0.001) were nominally significant. For urgency with frequency, only the IFG survived correction (aOR=11.45, 95% CI 3.10–42.33, *P*<0.001); ACC (aOR=11.53, *P*=0.002), age (aOR=1.08/year, *P*=0.002), and brain atrophy (aOR=2.86, *P*=0.019) were nominally significant. For isolated frequency, no predictor reached the Bonferroni threshold; the strongest was ACR (aOR=5.46, *P*=0.002), followed by male sex (aOR=10.62, *P*=0.024) and age (aOR=1.08/year, *P*=0.007).

**Figure 2.**
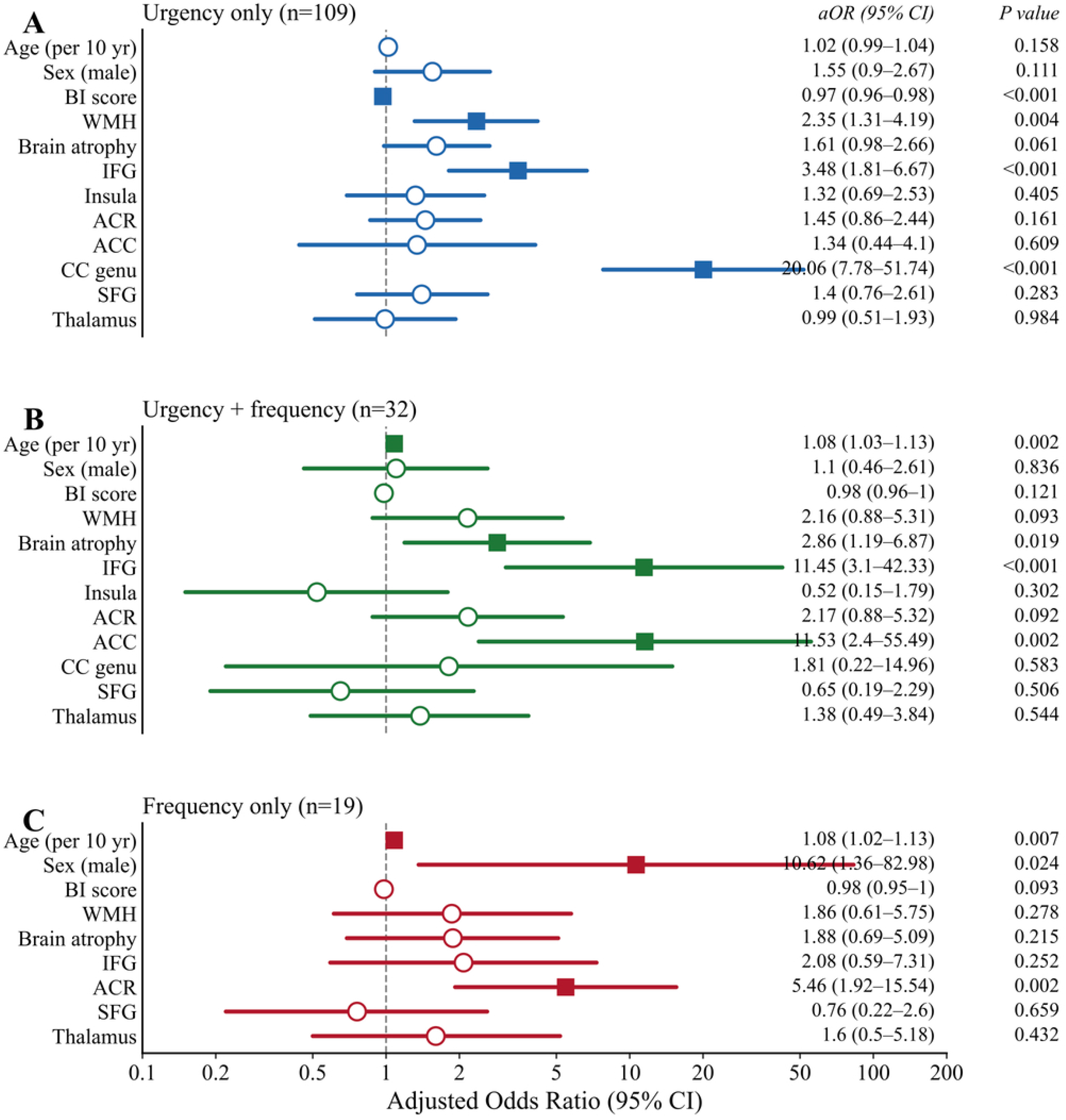
Multivariable logistic regression forest plot for three storage symptom subtypes. *P<0.05, **P<0.01, ***P<0.001 after Bonferroni correction.

Because of the small subgroup (n=19), the lesion-location model was limited to IFG and ACR. In an exploratory analysis, WMH≥3 and brain atrophy co-occurrence showed nominal univariable associations (Supplementary Table S3), but no significant atrophy×WMH interaction was found for any subtype (all interaction *P*>0.20).

### Side- Stratified Analysis and Laterality Patterns

Side-stratified analysis revealed divergent laterality profiles across subtypes (Table 2, Figure 3). For isolated urgency, laterality was structure-specific: right-hemisphere dominance was seen for the IFG (OR=5.54, 95% CI 2.25–13.62, *P*<0.001), whereas left-sided dominance was observed for the CC genu (OR=0.01, 95% CI 0.00–0.15, *P*<0.001; all 15 left-sided CC genu lesions involved isolated urgency), SFG (OR=0.12, *P*=0.021), and ACC (OR=0.13, *P*=0.018). This dissociation—right lateral-frontal and IFG yet left CC genu—indicates isolated urgency is not a simple right-hemisphere phenomenon.

**Figure 3.**
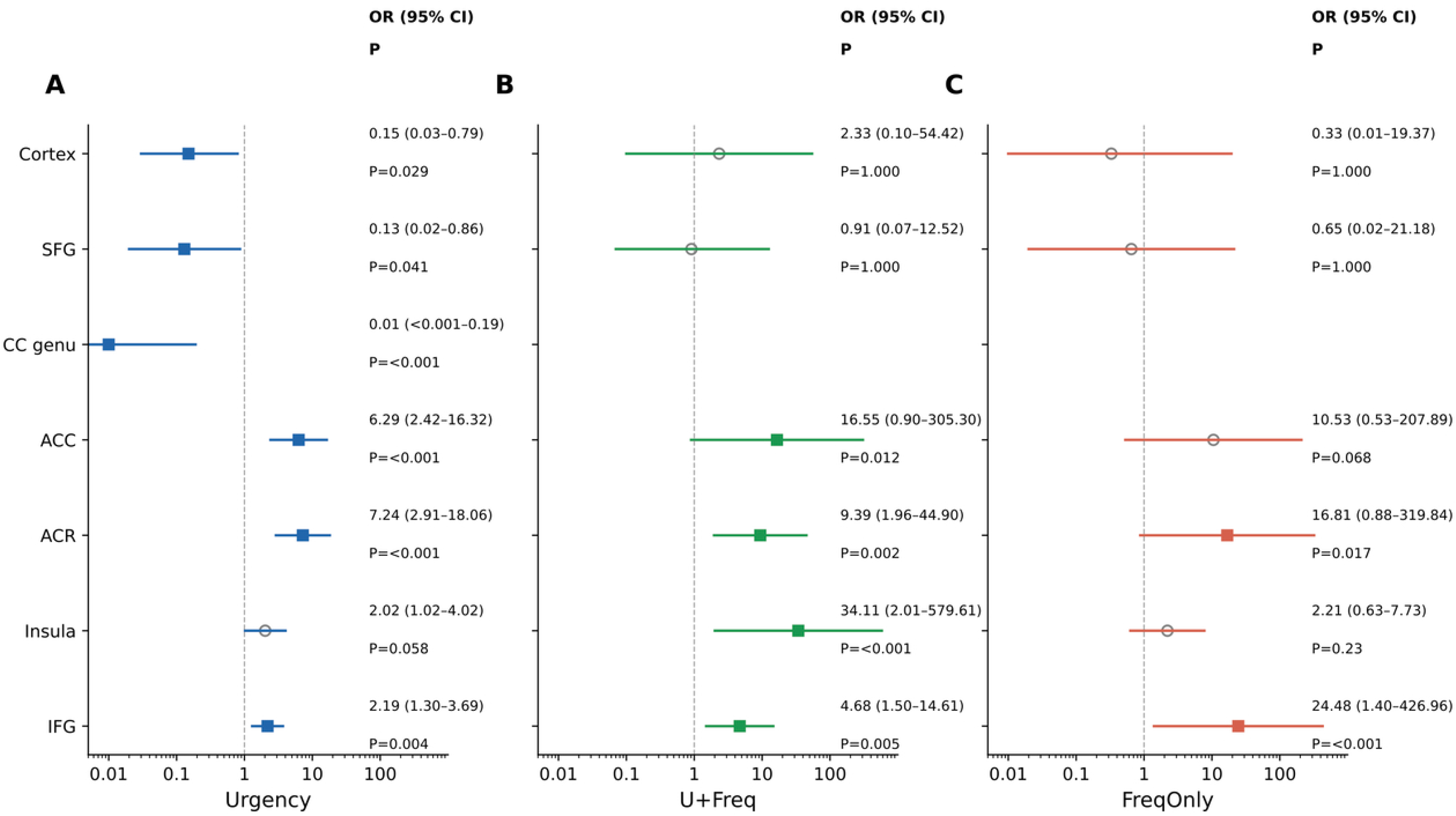
Side-stratified odds ratios of right vs. left hemisphere involvement across lesion locations for each symptom subtype (Table 2). Laterality odds ratios compare right-versus left-hemisphere involvement within each structure and reflect hemispheric asymmetry rather than causal strength.

**Table 2.** Side-stratified analysis of lesion laterality across symptom subtypes. Odds ratios compare right vs. left hemisphere involvement within each lesion location.

| Lesion location | Left n | Left Urgency only | Left U+Freq | Left Frequency only | Right n | Right Urgency only | Right U+Freq | Right Frequency only | Urgency only OR (95% CI) | Urgency only P | U+Freq OR (95% CI) | U+Freq P | Frequency only OR (95% CI) | Frequency only P |
| --- | --- | --- | --- | --- | --- | --- | --- | --- | --- | --- | --- | --- | --- | --- |
| IFG | 86 | 7 (8.1%) | 2 (2.3%) | 0 (0.0%) | 82 | 27 (32.9%) | 10 (12.2%) | 4 (4.9%) | 5.54 (2.25-13.62) | <0.001 | 5.83 (1.24-27.50) | 0.016 | 9.92 (0.53-187.17) | 0.055 |
| Insula | 89 | 6 (6.7%) | 0 (0.0%) | 0 (0.0%) | 88 | 25 (28.4%) | 5 (5.7%) | 3 (3.4%) | 5.49 (2.12-14.18) | <0.001 | 11.79 (0.64-216.53) | 0.029 | 7.33 (0.37-143.98) | 0.121 |
| ACR | 172 | 15 (8.7%) | 0 (0.0%) | 4 (2.3%) | 165 | 24 (14.5%) | 13 (7.9%) | 7 (4.2%) | 1.78 (0.90-3.53) | 0.125 | 30.54 (1.80-518.11) | <0.001 | 1.86 (0.53-6.48) | 0.371 |
| ACC | 10 | 7 (70.0%) | 1 (10.0%) | 0 (0.0%) | 22 | 5 (22.7%) | 5 (22.7%) | 1 (4.5%) | 0.13 (0.02-0.68) | 0.018 | 2.65 (0.27-26.25) | 0.637 | 1.47 (0.05-39.12) | 1.000 |
| CC genu | 15 | 15 (100.0%) | 0 (0.0%) | 0 (0.0%) | 18 | 3 (16.7%) | 3 (16.7%) | 0 (0.0%) | 0.01 (0.00-0.15) | <0.001 | 7.00 (0.33-147.18) | 0.233 | — | 1.000 |
| SFG | 12 | 8 (66.7%) | 0 (0.0%) | 0 (0.0%) | 20 | 4 (20.0%) | 3 (15.0%) | 0 (0.0%) | 0.12 (0.02-0.63) | 0.021 | 5.00 (0.24-105.67) | 0.274 | — | 1.000 |

\*Abbreviations: IFG, inferior frontal gyrus; ACR, anterior corona radiata; ACC, anterior cingulate cortex; CC, corpus callosum; SFG, superior frontal gyrus; U, isolated urgency; UF, urgency with frequency; F, frequency only.
†OR (95% CI) and P from Fisher's exact test (right vs. left) in strictly unilateral lesions (n=1,440). CC genu OR<1 reflects left-side dominance. EM dash (—) denotes complete separation. \*P<0.05, \*\*P<0.01, \*\*\*P<0.001

In contrast, urgency with frequency exhibited diffuse right-hemisphere dominance across multiple structures: ACR (right vs. left OR=30.54, *P*<0.001), insula (OR=11.79, *P*=0.029), and IFG (OR=5.83, *P*=0.016). The extreme ACR and large insula values reflect left-sided case absence (zero cases, Haldane-corrected) rather than causal strength. In multivariable analysis, only the IFG was independently associated (aOR=11.45, *P*<0.001); ACR was not (aOR=2.17, *P*=0.092). This diffuse right-lateralized pattern distinguishes urgency with frequency from the more focal laterality of isolated urgency.

### Synergistic and Structure-Specific Effects

Right IFG and insula showed synergistic associations (Table 3; Figure 4A; Supplementary Table S2). Isolated IFG was associated with isolated urgency (OR=2.67, *P*=0.008) and more strongly with urgency with frequency (OR=9.43, *P*<0.001), whereas isolated insula showed no significant association (OR=0.92, *P*=0.407). Combined IFG+insula involvement conferred markedly higher risk of isolated urgency (OR=21.46, 95% CI 10.49–43.88, *P*<0.001; present in 42.3% of right-sided urgency patients vs. 3.3% of controls) and was also associated with urgency with frequency (OR=6.50, *P*=0.003) and isolated frequency (OR=9.75, *P*=0.008). The positive RERI suggests additive synergy, though wide confidence intervals indicate the signal is suggestive rather than definitive.

**Figure 4.**
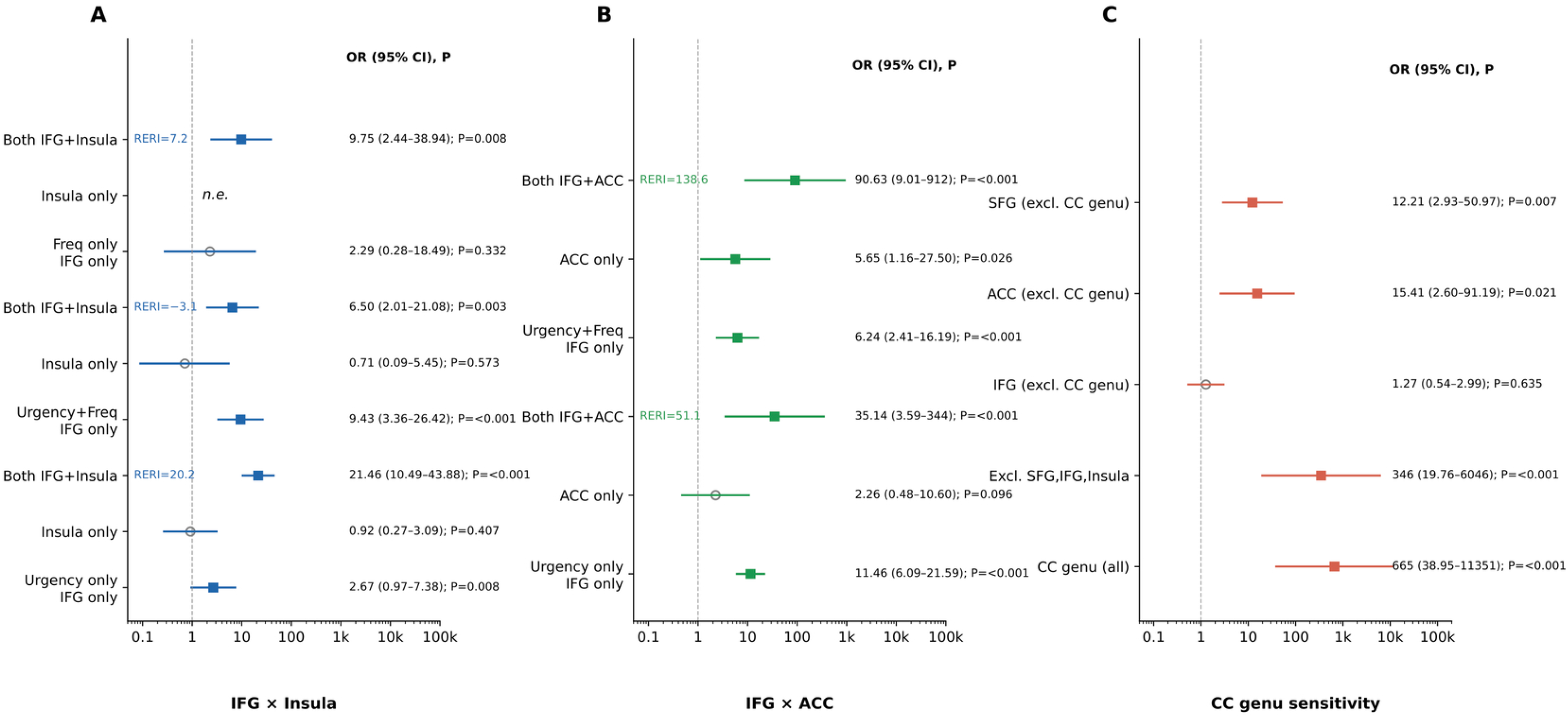
Right-sided inferior frontal gyrus (IFG) and insula synergy (Panel A), right-sided IFG and anterior cingulate cortex (ACC) co-involvement (Panel B), and left-sided genu of the corpus callosum (CC genu) specificity with sensitivity analysis (Panel C).

**Table 3.** Synergistic association of right-sided IFG and insula co-involvement, right-sided IFG and ACC co-involvement with post-stroke storage symptoms.

| Lesion pattern | n/N (%) | OR (95% CI) | P Value | Symptom subtype |
| --- | --- | --- | --- | --- |
| *Right-sided IFG and insula co-involvement (vs. right-sided normal controls)\** |  |  |  |  |
| IFG only | 5/52 (9.6) | 2.67 (0.97–7.38) | 0.008 | Urgency only |
| Insula only | 3/52 (5.8) | 0.92 (0.27–3.09) | 0.407 | Urgency only |
| Both IFG and insula | 22/52 (42.3) | 21.46(10.49–43.88) | <0.001 | Urgency only |
| IFG only | 6/22 (27.3) | 9.43 (3.36–26.42) | <0.001 | Urgency + frequency |
| Both IFG and insula | 4/22 (18.2) | 6.50(2.01–21.08) | 0.003 | Urgency + frequency |
| *Right-sided IFG and ACC co-involvement (vs. right-sided normal controls)\** |  |  |  |  |
| IFG only | 24/52 (46.2) | 11.46(6.09–21.59) | <0.001 | Urgency only |
| Both IFG and ACC | 3/52 (5.8) | 35.14(3.59–344.25) | <0.001 | Urgency only |
| IFG only | 7/22 (31.8) | 6.24(2.41–16.19) | <0.001 | Urgency + frequency |
| ACC only | 2/22 (9.1) | 5.65 (1.16–27.50) | 0.026 | Urgency + frequency |
| Both IFG and ACC | 3/22 (13.6) | 90.63(9.01–912.06) | <0.001 | Urgency + frequency |
| *Left-sided CC genu specificity for isolated urgency (vs. left-sided normal controls)†* |  |  |  |  |
| CC genu (all left-sided) | 15/15 (100) | 664.88(38.95–11350.85) | <0.001 | Isolated urgency |
| CC genu excluding SFG, IFG, insula | 9/9 (100) | 345.60(19.76–6046.00) | <0.001 | Isolated urgency |
| IFG excluding CC genu | 6/85 (7.1) | 1.27 (0.54–2.99) | 0.635 | Isolated urgency |
| ACC excluding CC genu | 2/5 (40.0) | 15.41 (2.60–91.19) | 0.021 | Isolated urgency |
| SFG excluding CC genu | 3/7 (42.9) | 12.21 (2.93–50.97) | 0.007 | Isolated urgency |

\*Abbreviations: ACC, anterior cingulate cortex; CC, corpus callosum; CI, confidence interval; IFG, inferior frontal gyrus; OR, odds ratio; SFG, superior frontal gyrus.
†Data are OR (95% CI) from logistic regression adjusted for age and sex; P from Wald test. \*P<0.05, \*\*P<0.01, \*\*\*P<0.001. Bold rows denote significant synergies (RERI>0, CI>0).

Right IFG and ACC showed a distinct pattern preferentially associated with urgency with frequency (Table 3, Figure 4B). Isolated IFG associated with both isolated urgency (OR=11.46, *P*<0.001) and urgency with frequency (OR=6.24, *P*<0.001), while isolated ACC was associated with urgency with frequency (OR=5.65, *P*=0.026) but not isolated urgency. Combined IFG+ACC involvement was preferentially associated with urgency with frequency (3/22 vs. 1/575 controls; Fisher *P*<0.001) and also with isolated urgency (3/52 vs. 1/575; Fisher *P*=0.002). This differential synergy—IFG+insula for isolated urgency versus IFG+ACC for urgency with frequency—suggests the specific combination of disrupted structures determines clinical phenotype, though the small number of co-involved cases warrants caution.

Left CC genu involvement showed complete specificity for isolated urgency (Table 3, Figure 4C): all 15 patients (100%; exact 95% CI, 78.2%–100%) with left CC genu lesions presented with isolated urgency (Fisher *P*<0.001). This specificity persisted after excluding concurrent frontal cortical involvement (9/9, 100%; exact 95% CI, 66.4%–100%; Fisher *P*<0.001). Of these 9, 3 were left-handed with right CC genu lesions, confirming the critical substrate is the dominant-hemisphere callosal pathway rather than anatomical left side. In contrast, right CC genu produced urgency in only 3/18 patients (16.7%). Left IFG without CC genu rarely elicited urgency (6/85, 7.1%; OR=1.27, *P*=0.635), while left ACC and SFG without CC genu showed moderate associations (40.0% and 42.9%). This hierarchy—CC genu > medial frontal > lateral frontal—indicates the interhemispheric callosal pathway plays a primary role in bladder inhibitory control. Clinical follow-up of CC genu patients showed that all 15 had complete resolution of urinary urgency within one month Thalamus, basal ganglia, and cerebellum showed no significant positive associations with any symptom subtype; brainstem was not independently associated in multivariable analysis.

A dose–response analysis of the cumulative number of affected key structures (CC genu, SFG, ACC, IFG, insula, ACR) in the ipsilateral hemisphere confirmed the marked right-hemisphere lateralization in strictly unilateral lesions (Figure 5A). In right-sided lesions, urgency prevalence rose steeply and monotonically from 4.5% (0 structures) to 9.9%, 26.5%, and 60.0% (1, 2, ≥3 structures). Left-sided lesions showed no consistent gradient (3.3%, 12.0%, 20.8%, 17.6%). This right-lateralized monotonic dose–response indicates cumulative right-hemisphere structural burden is a key determinant of storage symptoms; left-sided urgency instead relies on disruption of the left CC genu callosal pathway that conveys frontal inhibitory signals (Figure 3; Supplementary Table S2).

**Figure 5.**
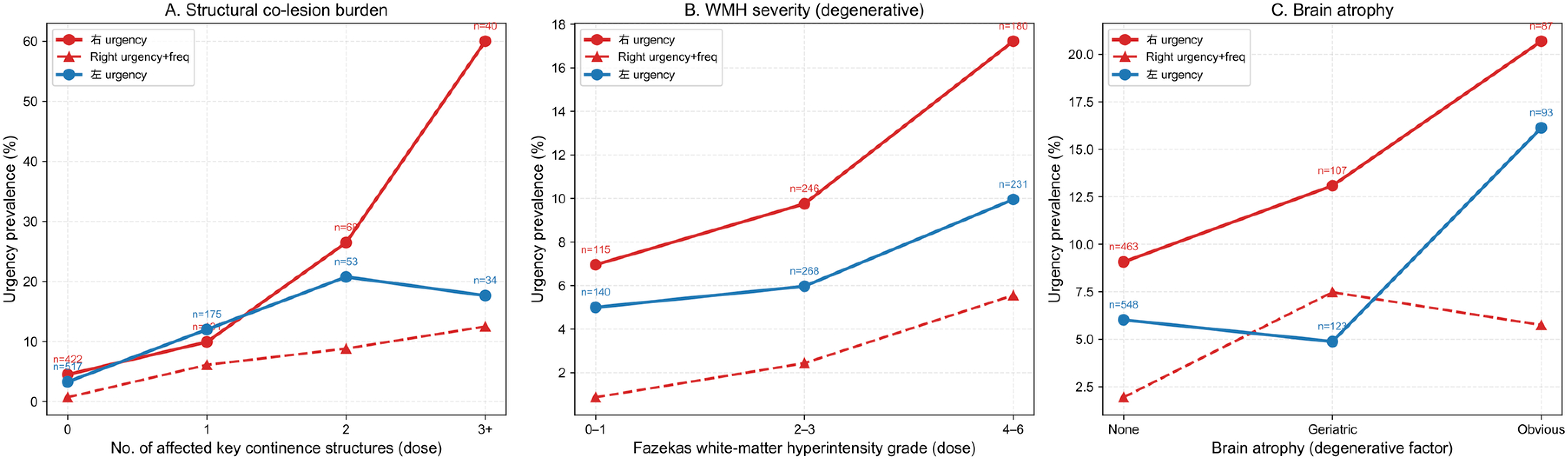
Dose–response relationship between the number of affected key continence structures and the severity of cerebrovascular burden on storage symptoms (data from Figure 5). (A) Prevalence of storage urgency by the cumulative number of affected key continence structures (CC genu → SFG → ACC → IFG → insula → ACR) in the ipsilateral hemisphere (dose, 0/1/2/3+), by side. A steep monotonic gradient is present for right-sided lesions but not left-sided lesions; left-sided urgency is instead associated specifically with involvement of the left CC genu, a pivotal commissural hub for the urgency signal (Figure 3; Supplementary Table S2). (B) Prevalence of storage urgency by white-matter hyperintensity severity (Fazekas grade 0–1/2– 3/4–6). (C) Prevalence of storage urgency by brain atrophy (none / geriatric change / obvious). Solid lines, overall urgency (right, red; left, blue); dashed red line, right-sided urgency with frequency.

## DISCUSSION

This study identifies three mechanistically distinct post-stroke storage symptom subtypes— isolated urgency, urgency with frequency, and isolated frequency—each defined by unique neural correlates, lateralization patterns, and clinical risk profiles. In contrast to the traditional unitary OAB framework of suprapontine disinhibition,^7–8,11^ these three phenotypes arise from separate lesion networks with separable neural substrates. The largely distinct independent predictors across subtypes (Figure 2) support a discrete pathway model rather than a continuum.

The known anatomy of supraspinal lower urinary tract control explains how pathway-specific lesion patterns produce distinct symptom profiles. Comparable prevalence across ischemic and hemorrhagic etiologies suggests generalizability of these subtype-specific neural mechanisms.

### Isolated Urgency: A Bilateral but Asymmetric Disinhibition Network

Isolated urgency after stroke engages a bilateral yet structurally asymmetric network. At the hemispheric level, no overall lateralization was observed (right hemisphere OR =1.00, P =0.992), a finding that challenges the notion of consistent right-hemispheric dominance in post-stroke micturition disturbance, a subject of ongoing debate.^8^ Earlier stdies d id not distinguish isolated urgency from urgency with frequency, potentially conflating mechanistically distinct subtypes. Structure-level analysis clarifies this paradox: the left genu of the corpus callosum was exclusively associated with isolated urgency, whereas the right inferior frontal gyrus alone demonstrated a strong and specific association with this subtype; this effect was further amplified by synergistic interaction with the right insula.

The left CC genu showed high specificity for isolated urgency, persisting after excluding concurrent frontal cortical damage (9/9, 100%; 95% CI, 66.4%–100%), a pattern absent for any single frontal region. Notably, the left SFG and ACC showed high frequencies of co-involvement in patients with isolated urgency (SFG: 4/9, 44.4%; ACC: 3/9, 33.3%), yet neither achieved the complete specificity of the left CC genu. This suggests that while the left SFG and ACC participate in the frontal inhibitory network for micturition control, they are not individually sufficient to produce isolated urgency upon lesion; rather, their disruption contributes to urgency through disconnection of the CC genu as the common convergence white-matter pathway. The left superior frontal lobe has been identified as the primary cortical site for volitional suppression of micturition,^16^ with its descending inhibitory signals crossing through the genu of the corpus callosum to reach right-hemisphere efferent pathways. Right CC genu lesions rarely caused isolated urgency (16.7%), supporting a transcallosal route—left frontal inhibitory signals traverse the left CC genu and reach the right hemisphere through distinct white matter pathways. The rapid resolution of urgency in patients with CC genu involvement suggests swift establishment of compensatory pathways.

The right IFG was the sole cortical region surviving strict Bonferroni correction (aOR=3.48, P<0.001), establishing it as the most robust cortical predictor of isolated urgency. A similar IFG-driven inhibitory network has been proposed for the lower urinary tract based on structural imaging in urgency incontinence,^3,17^ and our lesion data provide direct causal evidence for this framework. This finding revises the classical model emphasizing the insula as the primary cortical relay for bladder afferent signals,^10,18^ with the IFG receiving comparatively little attention in continence literature. Our data show that isolated insular lesions did not associate with isolated urgency, contradicting prior reports that the insula is the main driver of OAB. Right IFG–insula synergy substantially amplified urgency risk (OR=21.46), implying that disruption of a right-lateralized sensory–motor integration circuit—rather than right insula damage alone— most severely impairs voluntary continence.

### Urgency with Frequency: Right-Hemisphere Dominance and Cumulative Dose-Response

In contrast, U+Freq exhibited robust right-hemisphere dominance. However, within the multivariable model, the IFG was the sole independent predictor(aOR=11.45), whereas the ACR and ACC failed to reach significance. This indicates that the right-lateralized cortical command node — rather than subcortical white-matter tracts alone — is the principal driver of this phenotype. This suggests a distributed network disruption where right-hemisphere cortical centers and their projections are compromised, consistent with prior lesion studies linking right-sided strokes to urgency and frequency.^18^

The involvement of the ACC in U+Freq aligns with its role in interoceptive processing and discomfort monitoring. As part of the salience network, the ACC–IFG circuit filters behaviorally relevant internal signals, including bladder afferents.^11^ While asymptomatic controls exhibit sensory gating that prevents low-volume afferents from reaching conscious awareness,^19^ U+Freq patients likely suffer from corrupted gating due to ACC damage. This dysfunction lowers the sensory threshold, causing urgency at small volumes, while concurrent IFG involvement impairs the suppression of the micturition response. Our data confirm a specific synergistic interaction between the right IFG and ACC in U+Freq, which was significantly stronger than in isolated urgency (Table 3, Figure 4B). This distinct IFG+ACC synergy suggests that ACC-mediated interoceptive dysregulation is the key mechanism converting simple urgency into the full OAB complex, consistent with the known predominance of right-hemisphere lesions in urinary frequency and urgency.^20^

The U+Freq phenotype was underpinned by an aging brain substrate: in the multivariable model, brain atrophy was an independent predictor (aOR=2.86, P=0.019) together with increasing age (P=0.002), whereas the high WMH burden descriptively present in this subtype (59.4%) did not retain independent significance (aOR=2.16, 95% CI 0.88–5.31, P=0.093). White matter hyperintensities have been proposed to constitute a brain disease underlying OAB in the elderly by preferentially disrupting frontal subcortical white matter tracts.^20,21^ Wakefield et al. (2010) demonstrated that WMH volume predicted functional decline in voiding among community-dwelling older adults,²¹ and Kuchel et al. (2009) demonstrated that right-sided ACR WMH specifically predicted urinary incontinence severity.^20^ The convergence of acute right-hemisphere stroke with chronic small vessel disease and brain atrophy in U+Freq patients represents a ’double hit’—acute disruption of cortical command nodes superimposed on compromised white matter connectivity in an aging brain—producing the most clinically severe phenotype.

### Isolated Frequency: White Matter Disconnection Pathway

Isolated frequency presented a distinct and mechanistically informative profile: ACR involvement was the strongest independent predictor (aOR=5.46, 95% CI 1.92–15.54, *P*=0.002), with no significant cortical or cingulate involvement, although this association did not reach the conservative Bonferroni threshold. This supports a ’white matter disconnection’ subtype in which damage to ascending and descending fibers in the ACR disrupts normal afferent bladder sensory processing without triggering the cortical motor urgency response.

The ACR contains thalamocortical and corticothalamic projections critical for relaying bladder filling sensations.^7^ Disruption of these fibers may alter perception of bladder distension, producing increased frequency through afferent sensitization rather than detrusor overactivity. The striking male predominance (94.7%, P=0.035) and oldest median age (72 years) support a chronic cerebrovascular mechanism. This triad—advanced age, male sex, and ACR white matter disruption—recapitulates the risk profile of cerebral small vessel disease, which disproportionately affects men and preferentially involves periventricular and deep white matter.^16,20^ The male predominance diverges sharply from idiopathic OAB, which is more prevalent in women,²² underscoring that post-stroke isolated frequency is not OAB without urgency but a distinct entity driven by vascular white matter injury.

This white-matter-predominant profile aligns with Gelber et al.’s (1993) three-mechanism framework for post-stroke incontinence^23^ but extends it by identifying a fourth pathway that symptom subtyping revealed. Our findings also resonate with the LADIS study, which demonstrated that WMH independently contribute to LUTS beyond age and vascular risk factors,^24^ confirming that this relationship holds even in the post-stroke setting. Age acts as a primary confounder driving both neuroimaging markers and urinary symptoms; the apparent relationship between global structural damage and LUTS reflects shared aging burden rather than a direct causal pathway. Once age is accounted for, the specific topography of white matter injury—not the global burden of atrophy or WMH—determines the clinical phenotype. This age-confounding effect is particularly relevant to the LADIS study, which enrolled elderly individuals (aged 65–84 years) in whom age and WMH burden are inherently collinear.^24^ ***Subacute Phase as a Window to the Normal Human Urinary Control Network***

The subacute stroke population, assessed within the first month after onset, offers a unique window in which symptom-lesion associations reflect the functional necessity of regions for normal storage control rather than post-recovery residual deficits. Three observations support this view. First, the near-complete specificity of left CC genu involvement for isolated urgency (15/15, 100%) identifies a callosal pathway indispensable for interhemispheric coordination of bladder inhibition. Second, the right IFG-insula synergy— neither structure alone produced urgency but co-involvement yielded a >20-fold risk (OR=21.46) — indicates that both interoceptive and executive-inhibitory nodes must be simultaneously disrupted; compensation from the intact hemisphere is insufficient when both components are compromised. Third, the absence of significant associations with brainstem, thalamus, or basal ganglia indicates that these phylogenetically older regions have robust bilateral redundancy, whereas the identified cortical and white matter nodes have limited redundancy, making them rate-limiting for normal storage. Notably, urgency associated with left CC genu involvement resolved rapidly, often within days, reflecting transient interhemispheric disruption rather than fixed damage. Accompanying frequency tended to persist longer.

### Relationship to Existing Models and Therapeutic Implications

Our findings extend the suprapontine disinhibition model^7,25^ with neuroanatomical granularity. The framework reconciles inconsistent laterality findings across studies^2,8,26^: overall hemispheric effects are confounded by subtype-specific lateralization patterns that cancel at the aggregate level. This directly addresses Geerling et al.’s (2020) call for a sufficiently powered lesion-symptom mapping studyto test the non-dominant hemisphere’s role in micturition.^9^

The present data argue against the prevailing view that post-storage symptoms are independent of lesion site.¹³,²¹ Although aggregate hemispheric side was uninformative, specific lesion location was the dominant determinant of subtype: left CC genu produced isolated urgency with near-complete specificity; right frontal and cingulate lesions produced urgency with frequency; and anterior corona radiata disconnection produced isolated frequency. Global factors did not erase this site specificity but loaded differentially across subtypes. The persistence of a triple dissociation after adjustment for these factors indicates that lesion location, not the aggregate burden of chronic cerebrovascular disease, dictates the storage phenotype. Stroke etiology (ischemic vs hemorrhagic) was not an independent predictor in any subtype.

These findings constitute a paradigm shift from the insula-centered model of supraspinal bladder control toward an IFG-anchored, subtype-specific framework. Our lesion-mapping data (n=1,498) demonstrate that the right IFG, rather than the insula, is the pivotal causal node for isolated urgency — a discrepancy explained by the difference between functional imaging^27^(regional activation, potentially epiphenomenal) and lesion-symptom mapping (causal necessity). The right IFG is a well-established causal node for response inhibition,^14,15^ and its lesion produces a deficit directly paralleling the core symptom of inability to suppress urgency.

These three dissociable phenotypes unify within a single neuroanatomical framework. Bladder afferent information ascends bilaterally, relays through the right anterior insula,^10,11,15^ and converges on the right IFG, which—together with the anterior cingulate cortex—forms the right-lateralized sensorimotor integration loop for urgency suppression.^14,20,28^ Right IFG-insula co-lesion exhibited synergistic enhancement beyond additive effects (Table 3). The left hemisphere generates inhibitory commands through a distributed prefrontal-cingulate network; no single left cortical locus alone produced urgency. All left-hemisphere inhibitory signals must converge on the left CC genu to reach contralateral targets, making this callosal conduit the unique bottleneck whose interruption releases the pontine micturition center from cortical suppression. Disruption of the ACR, which carries ascending and descending bladder projections,^7^ alters perception of bladder distension and increases frequency through afferent sensitization rather than detrusor overactivity.

These dissociable pathways carry direct therapeutic implications. Isolated urgency (left CC genu + right IFG-insula) may respond to antimuscarinics combined with urge suppression. U+Freq (fronto-cingulate disruption with brain atrophy) requires vascular risk factor optimization with cognitive-behavioral scheduling. Isolated frequency (ACR disconnection) benefits from sensory-level neuromodulation—percutaneous tibial nerve or sacral neuromodulation.^29^ Based on these substrates, we have initiated a pilot protocol of navigated rTMS targeting each subtype’s network. Lesion-guided stratification may complement standard OAB management and, by alleviating urgency-driven fluid restriction, potentially reduce recurrent cerebrovascular events.

### Aphasia Does Not Impair Identification of Storage Symptoms: An Imperative for Early Rehabilitation

Aphasia was not a barrier to identifying post-stroke storage symptoms. Overall OR=0.51 (P=0.081), with prevalence declining from 9.3% in asymptomatic controls to 0% in urgency with frequency and isolated frequency (Cochran-Armitage Z=-2.26, P=0.024). This challenges the earlier assumption that aphasia obscures or inflates symptom ascertainment.^2^ Rather than masking symptoms, aphasia carried a numerically inverse relationship, consistent with the largely right-lateralized lesion networks underlying storage phenotypes.

Our structured bedside strategy operationalized urgency through non-verbal cues (restlessness, bed-tapping, gesturing to void), corroborated by continuous caregiver observation of voiding episodes. The absence of systematic under-reporting confirms that storage symptoms are clinically decipherable even without expressive language. This reframes aphasia from an obstacle into a structured communication task: training patients to signal bladder need and caregivers to recognize these cues can preserve continence, reduce catheter dependence, and improve quality of life during the subacute period.

### Limitations

Several limitations should be acknowledged. First, symptom classification relied on clinical assessment rather than urodynamic confirmation.¹ However, our population comprised exclusively subacute stroke patients assessed within 30 days of onset, a window in which behavioral symptoms closely reflect acute lesion effects before compensatory mechanisms or secondary neuroplasticity obscure the primary pathology. Moreover, structured bedside protocols incorporating non-verbal cues and caregiver reports have demonstrated validity in prior stroke populations,^28^ and our aphasia findings (no association with storage symptoms) argue against systematic misclassification.

Second, the small sample sizes in U+Freq (n=32) and FreqOnly (n=19) limit statistical power, reflected in wide confidence intervals. Left-sided U+Freq was extremely rare (7/779, 0.9%), precluding reliable statistical inference; these data are presented descriptively only. Future multicenter collaborations will be needed to adequately characterize this phenotype.

Third, lesion mapping relied on clinical radiological reports rather than voxel-based lesion-symptom mapping (VLSM). This approach has complementary strengths: the anatomical structures we analyzed can be reliably identified on routine clinical MRI, enhancing translational applicability. However, VLSM might reveal additional subtle associations at the voxel level that our region-of-interest approach could miss.

Fourth, the scope of the present study is deliberately confined to storage-phase symptoms (urgency and frequency) after stroke. Voiding-phase symptoms (hesitancy, weak stream, incomplete emptying) were not systematically assessed and may have distinct neural substrates. Future work should integrate both storage and voiding domains to provide a comprehensive map of post-stroke lower urinary tract dysfunction.

## Conclusions

Storage symptoms after stroke comprise at least three mechanistically distinct subtypes with largely distinct neural correlates. Isolated urgency arises from left CC genu (interhemispheric disinhibition) and right IFG-insula synergy (sensory-motor pathway). Urgency with frequency reflects right IFG-ACC synergistic disruption within a broader fronto-cingulate-ACR network superimposed on chronic cerebrovascular disease. Isolated frequency without urgency represents a white matter disconnection syndrome driven by ACR damage. These findings challenge the unitary OAB paradigm and provide a neuroanatomically grounded subclassification of post-stroke storage symptoms, offering a theoretical foundation for subtype-stratified treatment selection.

## Author Contributions

Zhaoxia Wang: Conceptualization, Methodology, Formal Analysis, Investigation, Writing - Original Draft. Huixian Yu: Supervision, Writing – Review & Editing. Dawei Zang: Supervision. Zhike Yin: Investigation. Yumei Zhang, Pei Dai, Sihao Liu, Qian Wang, Yuexiu Li, Changbin Liu, and Ran Liu: Project Administration. All authors read and approved the final manuscript.

## Data Supplement

The online-only Data Supplement is available with this article.

Supplementary Table S1. Univariate analysis of lesion location across storage symptom subtypes.

Supplementary Table S2. Distributions of right-sided IFG/insula/ACC lesion patterns and detailed odds ratios across storage symptom subtypes.

Supplementary Table S3. Synergistic association between brain atrophy and white matter hyperintensity burden (Fazekas score ≥3) across post-stroke storage symptom subtypes.

## Data Availability

All relevant data are within the paper and its supplementary information files.

## References

[1] Pizzi A, Falsini C, Martini M, et al. Urinary incontinence after ischemic stroke: clinical and urodynamic studies. Neurourol Urodyn. 2014;33(4):420–425.

[2] Mehdi Z, Birns J, Bhalla A. Post-stroke urinary incontinence. Int J Clin Pract. 2013;67(11):1128–1137.

[3] McKenzie P, Badlani GH. The incidence and etiology of overactive bladder in patients after cerebrovascular accident. Curr Urol Rep. 2012;13(5):402–406.

[4] Abrams P, Cardozo L, Fall M, et al. The standardisation of terminology of lower urinary tract function: report from the Standardisation Sub-committee of the International Continence Society. Neurourol Urodyn. 2002;21(2):167–178.

[5] Kavia RBC, DasGupta R, Fowler CJ. Functional imaging and the central control of the bladder. J Comp Neurol. 2005;493(1):27–32.

[6] DasGupta R, Kavia RBC, Fowler CJ. Cerebral mechanisms and voiding function. BJU Int. 2007;99(4):731–734.

[7] Fowler CJ, Griffiths D, de Groat WC. The neural control of micturition. Nat Rev Neurosci. 2008;9(6):453–466.

[8] Sakakibara R, Hattori T, Yasuda K, Yamanishi T. Micturitional disturbance after acute hemispheric stroke: analysis of the lesion site by CT and MRI. J Neurol Sci. 1996;137(1):47–56.

[9] Tish MM, Geerling JC. The brain and the bladder: forebrain control of urinary (in)continence. Front Physiol. 2020;11:658.

[10] Griffiths D. Neural control of micturition in humans: a working model. Nat Rev Urol. 2015;12(12):695–705.

[11] Griffiths D, Tadic SD. Bladder control, urgency, and urge incontinence: evidence from functional brain imaging. Neurourol Urodyn. 2008;27(6):466–474.

[12] Michels L, Blok BFM, Gregorini F, et al. Supraspinal control of urine storage and micturition in men—an fMRI study. Cereb Cortex. 2015;25(10):3369–3380.

[13] Kim TG, Yoo KH, Jeon SH, Lee HL, Chang SG. Effect of dominant hemispheric stroke on detrusor function in patients with lower urinary tract symptoms. Int J Urol. 2010;17(7):656–660.

[14] Aron AR, Robbins TW, Poldrack RA. Inhibition and the right inferior frontal cortex: one decade on. Trends Cogn Sci. 2014;18(4):177–185.

[15] Critchley HD, Wiens S, Rotshtein P, et al. Neural systems supporting interoceptive awareness. Nat Neurosci. 2004;7(2):189–195.

[16] Fazekas F, Chawluk JB, Alavi A, et al. MR signal abnormalities at 1.5 T in Alzheimer’s dementia and normal aging. AJR Am J Roentgenol. 1987;149(2):351–356.

[17] Clarkson BD, Karim HT, Chermansky CJ, Banihashemi L, Tyagi S, Griffiths DJ, et al. Changes in brain response to urgency before and after treatment of urgency urinary incontinence with onabotulinumtoxin A. Neurourol Urodyn. 2022;41(8):1703–1710. doi:10.1002/nau.25012.

[18] Funayama M, Koreki A, Takata T, Nakagawa Y, Mimura M. Post-stroke urinary incontinence is associated with behavior control deficits and overactive bladder. Neuropsychologia. 2024 Aug 13;201:108955.

[19] Griffiths D, Derbyshire S, Stenger A, Resnick N. Brain control of normal and overactive bladder. J Urol. 2005;174(5):1862–1867.

[20] Kuchel GA, Moscufo N, Guttmann CR, et al. Localization of brain white matter hyperintensities and urinary incontinence in community-dwelling older adults. J Gerontol A Biol Sci Med Sci. 2009;64A(8):902–909.

[21] Wakefield DB, Moscufo N, Guttmann CR, et al. White matter hyperintensities predict functional decline in voiding, mobility and cognition in older persons. J Am Geriatr Soc. 2010;58(2):275–281.

[22] Griffiths D. Cerebral control of bladder function. Curr Urol Rep. 2004;5(5):348–352.

[23] Gelber DA, Good DC, Laven LJ, Verhulst SJ. Causes of urinary incontinence after acute hemispheric stroke. Stroke. 1993;24(3):378–382.

[24] Poggesi A, Pracucci G, Chabriat H, et al. Urinary complaints in nondisabled elderly people with age-related white matter changes: the Leukoaraiosis And DISability (LADIS) Study. J Am Geriatr Soc. 2008;56(9):1638–1644.

[25] Blok BF, Willemsen AT, Holstege G. A PET study on brain control of micturition in humans. Brain. 1997;120(1):111–121.

[26] Kuroiwa Y, Tohgi H, Ono S, Itoh M. Frequency and urgency of micturition in hemiplegic patients: relationship to hemisphere laterality of lesions. J Neurol. 1987;234(1):100–102.

[27] Kreydin EI, Abedi A, Morales L, Montero S, Kohli P, Ha N, et al. Neural mechanisms of poststroke urinary incontinence: results from an fMRI study. Stroke. 2025;56(6):1516–1527.

[28] Bou Kheir G, Verbakel I, Hervé F, Bauters W, Abou Karam A, Holm-Larsen T, et al. OAB supraspinal control network, transition with age, and effect of treatment: a systematic review. Neurourol Urodyn. 2022;41(6):1224–1239.

[29] Pericolini M, Finazzi Agrò E, Chesnel C, et al. Cortical, spinal, sacral, and peripheral neuromodulations as therapeutic approaches for the treatment of lower urinary tract symptoms in multiple sclerosis patients: a review. Neuromodulation. 2022;25(8):1065–1075.

